# Psychosis-like experiences and cognition in young adults: an observational and Mendelian randomisation study

**DOI:** 10.1101/2021.05.06.21256771

**Authors:** Caroline Skirrow, Steph Suddell, Liam Mahedy, Ian S. Penton-Voak, Marcus R. Munafò, Robyn E. Wootton

## Abstract

**Background:** Psychosis-like experiences (PLEs) are common and associated with mental health problems and poorer cognitive function. There is limited longitudinal research examining associations between cognition and PLEs in early adulthood.

**Aims:** We investigated the association of PLEs with different domains of cognitive function, using cross-sectional and longitudinal observational, and Mendelian randomisation (MR) analyses.

**Method:** Participants from the Avon Longitudinal Study of Parents and Children (ALSPAC) completed tasks of working memory at age 18 and 24, and tasks of response inhibition and facial emotion recognition at age 24. Semi-structured interviews at age 18 and 24 established presence of PLEs (none vs. suspected/definite). Cross-sectional and prospective regression analysis tested associations between PLEs and cognition (N=3,087 imputed sample). MR examined causal pathways between schizophrenia liability and cognition.

**Results:** The fully adjusted models indicated that PLEs were associated with poorer working memory performance (cross-sectional analyses: *b*=−0.18, 95% CI −0.27 to −0.08, p<0.001; prospective analyses: *b*=−0.18, 95% CI −0.31 to −0.06, p<0.01). A similar pattern of results was found for PLEs and response inhibition (cross-sectional analyses: *b*=7.29, 95% CI 0.96 to 13.62, p=0.02; prospective analyses: *b*=10.29, 95% CI 1.78 to 18.97, p=0.02). We did not find evidence to suggest an association between PLEs and facial emotion recognition. MR analyses were underpowered and did not support observational results.

**Conclusions:** In young adults, PLEs are associated with poorer concurrent and future working memory and response inhibition. Better powered genetically informed studies are needed to determine if these associations are causal.

---

Psychosis-like experiences (PLEs; subclinical delusions, hallucinations or other unusual experiences) are common in the general population. Around 10% of children and adolescents experience PLEs^1^, and among adults lifetime prevalence is estimated at 6%^2^. PLEs are more common in younger children, with rates decreasing across age. Longitudinal studies show predominantly remitting or intermittent trajectories, and a minority have persistent trajectories^3,4^. Although common in early life, onset of PLEs may occur throughout the lifespan^2^. Early PLEs enhance the risk not only for later developing psychotic disorders, but also for other mental health conditions, including anxiety, affective, behavioural and substance use disorders^1^, and broader functional and social deficits^5^.

For individuals who subsequently develop schizophrenia, meta-analyses show that generalised cognitive deficits are typically present by age 16^6^, and that reductions in IQ over time increase the risk of schizophrenia diagnosis^7^. Cognitive impairments including intellectual functioning, working memory, verbal and visual memory, executive function and attention, are present in individuals at high risk of developing schizophrenia before onset of psychotic illness^8^. Research shows limited evidence of decline during, or after, the first episode of psychosis^9^. Together, evidence supports a neurodevelopmental rather than neurodegenerative model of schizophrenia. Longitudinal studies are important for revealing emergence and trajectory of these features over time^8^.

However, how cognitive functions develop in the context of PLEs is less understood. Meta-analysis shows moderate deficits in IQ in individuals with PLEs, alongside more modest impairments in memory, working memory, and processing speed^10^. A recent study in the Avon Longitudinal Study of Parents and Children (ALSPAC), a large UK birth cohort, comparing healthy subjects, young people with psychotic disorder, and young people with PLEs suggested that only individuals with psychotic disorders at age 18 showed progressive IQ declines prior to onset of psychosis. Cognitive function of subjects with PLEs was intermediate between healthy subjects and those with psychotic disorder, and did not decline with age^11^. Evidence from an earlier study in ALSPAC showed that below average IQ at age 8 was associated with PLEs at age 12^12^. Poorer cognitive function may therefore predate the onset of PLEs as it does for psychotic disorders.

A longer-term developmental lens is needed to further our understanding of the relationship between PLEs, psychotic illness and cognition across the lifespan. As young people reach and progress through early adulthood, cognitive function in specific domains reach maturity^13^. This is also a time of vulnerability for development of PLEs^2^ and psychotic disorders^14^. To understand the relationship between cognition and PLEs it is therefore not sufficient to rely on data from childhood and adolescence as many functions continue to develop in young adulthood.

In the current study we used data from ALSPAC to investigate the relationship between PLEs and three cognitive domains in early adulthood: working memory, response inhibition, and emotion recognition. We aimed to: (1) examine the cross-sectional association between PLEs and cognition at age 24, (2) use longitudinal data at age 18 and 24 to test whether PLEs simply co-occur with poorer cognitive function or if they are preceded by poorer cognitive function, and (3) use Mendelian randomisation to explore causal effects between genetic liability for schizophrenia and cognitive impairment.

## Method

### Participants

ALSPAC (www.alspac.bris.ac.uk) is a birth cohort initially comprising 14,541 pregnancies in the former County of Avon, currently the Bristol area of the UK, with an estimated delivery date between April 1991 and December 1992. The parents completed regular postal questionnaires on aspects of the children’s development since birth, and from mid childhood (age 7.5 and above) children attended in-clinic assessments where they take part in a range of face-to-face interviews and tests. ALSPAC includes a wide range of phenotypic and environmental measures, genetic information and linkage to health and administrative records.

Detailed information about ALSPAC is available online www.bris.ac.uk/alspac and in cohort profiles^15–17^. A detailed overview of the study population that completed cognitive assessments at age 24, including attrition at the different measurement occasions is presented by Mahedy and colleagues^18^. Please note that the study website contains details of all the data that is available through a fully searchable data dictionary and variable search tool: http://www.bristol.ac.uk/alspac/researchers/our-data/.

### Ethics statement

Ethics approval for the study was obtained from the ALSPAC Ethics and Law Committee and the Local Research Ethics Committees. Written informed consent was obtained for the use of data collected via questionnaires and clinics from parents and participants following recommendations of the ALSPAC Ethics and Law Committee at the time. Consent for biological samples has been collected in accordance with the Human Tissue Act (2004). More details on ethics committees/institutional review boards are provided here: http://www.bristol.ac.uk/alspac/researchers/research-ethics/.

### Measures

Supplementary Figure S1 shows a timeline of the data used in analyses. Study data gathered from age 22 years onwards was collected using REDCap data capture tools^19^.

#### Psychosis-like Experiences (PLE)

PLEs were assessed via the psychosis-like symptoms interview (PLIKSi), a semi-structured interviewer-rated screening assessment. The interview lasts approximately 20 minutes and examines unusual experiences (unusual sensations, derealization, depersonalization, self-unfamiliarity, dysmorphophobia, partial object perception, and other perceptual abnormalities), and the three main domains of positive psychotic symptoms: hallucinations (auditory, visual, olfactory and tactile), delusions (being spied on, persecution, thoughts being read, reference, control, grandiose ability and other unspecified delusions) and bizarre symptoms (thought broadcasting, insertion and withdrawal).

Interviews were completed by trained psychology graduates who rated psychotic experiences as absent, suspected or definite. For each symptom, the interviewer read out a stem question from the interview schedule, and the participant responded with ‘yes’, ‘no’ or ‘maybe’. Where the participant responded ‘yes’ or ‘maybe’ the reply was then examined further with additional probes. Attribution questions identified if these were experienced only while falling asleep or waking, during high fever or under the influence of alcohol or other substances – in which case the experience was rated as not present. Symptoms were rated as definite where a clear example was provided.

For the purposes of this study, and in line with earlier research^12^, a binary variable was used for broad psychosis-like symptoms (none, versus suspected or definitely present). PLIKSi data was available from age 18 and 24. Symptom frequency in the last 6 months was examined at age 18 and symptom frequency in the last year was examined at age 24.

#### Cognitive assessments

Three computer-based cognitive assessments were delivered via E-Prime version 2 (Psychology Software Tools, pstnet.com).

##### Working memory

The N-back task^20^ is a widely used measure of working memory, requiring on-line monitoring, updating, and manipulation of remembered information^21^. We analysed data from the 2-back variant, administered at ages 18 and 24. Participants monitored a series of numbers (0-9), and indicated with a ‘1’ keystroke when the presented number was the same as the one presented 2 trials previously, and a ‘2’ keystroke when it was different. Stimuli were presented for 500ms, followed by a 3,000ms response window. Participants completed a practice block consisting of 12 trials containing two targets and response feedback, followed by an experimental block containing 48 trials with eight targets and no response feedback. The primary outcome measure was d-prime, the ratio of hits (correct detection of an n-back match) to false alarms (response during no match). Participants who responded to fewer than 50% of trials or with a negative d-prime were excluded. Higher d-prime scores index better working memory performance.

##### Response inhibition

A Stop Signal Task^22^ assessed the ability to prevent a prepotent motor response. Participants were instructed to respond rapidly to visual stimuli (an X or O) with the corresponding keys on a keyboard (X/O), unless they heard a tone (i.e., ‘stop signal) after the stimulus presentation. Targets were displayed for 1000ms, with a variable inter-stimulus interval of 500-100ms. Stop signal delay (SSD) was drawn from one of four adaptive staircases at 100ms, 200ms, 400ms or 500ms. On successful inhibition the staircase was adjusted by −25ms, and on failed inhibition it was adjusted by +25 (min SSD 25ms, max 800ms). Participants completed 32 practice trials, in which incorrect responses and slow responses were given the feedback “wrong” and “too slow”, respectively. This was followed by the task consisting of 4 blocks of 64 trials, with 25% randomly interspersed trials including a stop signal. The primary outcome measure was stop signal reaction time (SSRT), calculated as the difference between the median reaction time for go trials (Go Reaction Timemed) and an estimate of the median stop signal delay (SSRT = Go Reaction Timemed – Median Stop Signal Delay). Median Stop Signal Delay was the latency where each participant was likely to fail to inhibit 50% of trials. Lower SSRTs indicate better response inhibition.

##### Emotion recognition

A six alternative forced choice emotion recognition task (ERT) measured the ability to identify emotions in facial expressions^23^. Participants were presented with a series of face stimuli (male and female), each displaying one of six emotions: anger, disgust, fear, happiness, sadness, or surprise. They were instructed to click on the descriptor that best described the emotion. Images were displayed one at a time for 200ms, followed by a backwards mask of visual noise to prevent processing of afterimages for 250ms. Emotion intensity varied across 8 levels within each emotion from the prototypical emotion to an almost neutral face. Each individual stimulus was presented twice, giving a total of 96 trials with each emotion presented 16 times. Outcome measures included ERT hits (the total number of facial emotions accurately identified), and the number of correctly identified emotions for each discrete emotion category. Higher ERT scores indicate better performance.

#### Confounding variables

Background characteristics controlled for in analyses included participant sex (male, female), ethnicity (non-white, white), highest parental occupational level (grouped into four categories using the 1991 Office of Population Census and Statistics Classification^24^: unskilled or semiskilled manual, skilled manual or non-manual, managerial and technical, and professional), maternal education (categorised as: below O-level [examinations usually taken at age 16], O-level, above O-level), housing tenure (owned/mortgaged vs. other), maternal age at birth, maternal smoking in pregnancy (present, absent). Data was collected from questionnaire measures completed by parents in the antenatal and postnatal period.

Confounders relating to cognitive dysfunction included a history of head injury and a prior index of global cognitive function as measured by IQ. Head injury was defined as a cracked skull or unconsciousness at any time up to age 16 (present vs. absent), with data collected via questionnaires at age 4, 5, 6, 8, 11 years from parents and at age 16 years via self-report questionnaires. IQ was estimated from vocabulary and matrix reasoning subtests of the Wechsler Abbreviated Scale of Intelligence^25^ at age 15.

Mental health confounders included above threshold symptomatology for depression or anxiety on the Clinical Interview Schedule-Revised^26^ at both age 18 and age 24. Depression (mild, moderate or severe) and anxiety disorder (generalized anxiety disorder, social phobia, agoraphobia, and/or panic disorder) were coded as present or absent.

### Statistical methods

#### Observational analyses

Observational analyses were conducted in Stata (Release 15)^27^.

##### Study 1

Multivariable linear regression was used to examine the cross-sectional associations between PLEs and cognition (assessed at age 24). Summary variables from cognitive assessments (d-prime, SSRT, and ERT hits) were examined within one model. Individual ERT emotions (ERT hits for Anger, Disgust, Fear, Happy, Sad and Surprise) were entered into a second model to examine whether PLEs were associated with poorer recognition of specific emotions. A Wald test was used to test for differential effects across outcomes. Using complete case analysis without taking missing data into account can result in biased estimates^28^ so we imputed missing data using multivariate imputation by chained equations using the ‘ice’ package in Stata. Analyses were conducted on individuals with complete information on exposures, outcomes and confounders (n=2,134 for summary variables, n=2,300 for individual emotion ERT). Analysis of imputed data were based on 3,087 participants who provided data for all cognitive outcomes at age 24. A number of auxiliary variables known to be related to missingness were included in the imputation models and 100 data sets were imputed. An overview of predictors of attrition in ALSPAC participants completing cognitive testing at age 24 has been reported previously^18^.

##### Study 2

Multivariable linear regression was used to examine the prospective associations between PLEs and cognition. The association between PLEs assessed at age 18 and summary cognitive outcomes (d-prime, SSRT, and ERT hits) assessed at age 24 were examined in one model, and the association of PLEs at 18 with individual ERT emotions (ERT hits for Anger, Disgust, Fear, Happy, Sad and Surprise) at age 24 were entered into a second model, again using a Wald test to examine differential effects of exposure across outcomes. 1,788 individuals provided complete information on exposures, outcomes, and confounders for all summary cognitive outcomes, and 1,918 for all individual ERT emotions. Logistic regression was also used to examine the prospective association between working memory (d-prime) at age 18 and PLEs at age 24, for which 2,134 individuals provided complete information on exposures, outcomes, and confounders. Like Study 1, analysis of imputed data was based on 3,087 participants who provided data for all cognitive outcomes at age 24.

##### Model reporting

For both Study 1 and Study 2, we present unadjusted results, followed by results adjusted for: (i) sociodemographic variables (sex, ethnicity, parental occupation, maternal education, housing tenure, maternal age at birth and maternal smoking in pregnancy); (ii) additionally, confounders relating to cognitive dysfunction (history of head injury and IQ at age 15); and (iii) additionally, co-occurring depression and anxiety at age 24 in cross-sectional models, or depression and anxiety symptomatology at age 18 in prospective models. Models using multiply imputed data are presented as the main findings. Non-imputed (complete case) results are reported in the Online Supplement. Results from observational analyses are reported as unstandardized beta coefficients (*b*) or odds ratios (OR), as appropriate, both with 95% confidence intervals (95% CI).

#### Mendelian randomization

##### Study 3

Causal pathways between psychotic experiences and cognition were examined with Mendelian randomisation (MR) analyses. A genome-wide association study (GWAS) of PLEs across three adolescent population samples identified no genome-wide significant single nucleotide polymorphisms (SNPs) associated with PLE symptom domains, although genetic covariance between psychotic-like experience domains and schizophrenia genetic risk were documented^29^. For psychosis symptoms we therefore used 145-independent genome-wide significant loci associated with schizophrenia identified by Pardiñas and colleagues^30^ in a meta-analysis of genetic datasets including the CLOZUK sample, and an independent dataset from the Psychiatric Genomics Consortium (PGC) comparing 40,675 cases with schizophrenia and 64,643 controls^30^.

For cognition we used the full GWAS output in ALSPAC for each of our three primary cognitive outcome measures collected at age 24: d-prime, SSRT, and ERT hits, reported previously by Mahedy and colleagues^18^. GWAS of these cognitive measures yielded no statistically genome-wide significant SNPs which could serve as robust instrumental variables^31^. MR analysis was therefore conducted in only one direction, from schizophrenia liability as the exposure (using genome-wide significant SNPs for schizophrenia) to cognition as the outcome.

Two-sample MR analyses were carried out in the MR Base R package 0.4.26^32^ and compared four methods, each with different assumptions: (1) the inverse-variance weighted (IVW) method estimate, which is only accurate where all genetic variants are valid instrumental variables or when pleiotropy is balanced; (2) the weighted median, which allows for consistent estimation even when up to 50% of information comes from invalid instrumental variables^33^; (3) the mode-based estimate, which remains consistent when the largest number of similar individual-instrument causal effect estimates comes from valid instruments^34^; (4) MR Egger regression, which does not constrain the intercept to zero and therefore allows for directional pleiotropy in the instrumental variables^35^. Weighted median, mode-based and MR Egger methods provide useful sensitivity analyses in addition to the standard IVW approach, and consistent effects across all methods allows for more confident causal inference.

In one-sample MR analyses a polygenic risk score (PRS) for schizophrenia was calculated for each participant by computing the sum of risk alleles for Schizophrenia reaching genome-wide significance ^30^, weighted by the effect size estimate. Risk loci and the natural logarithm of odds ratios of effect for 131 out of 145 risk loci were entered into PLINK v.1.90^36^ to calculate the PRS, after excluding indels and palindromic loci. A logistic regression model was then used, regressing risk for schizophrenia on the presence of PLEs at age 24, to examine internal validity. Following this internal validation, the aim was to perform an instrumental variable regression, regressing the residuals from the PRS to PLE analysis onto cognitive outcome measures in the ALSPAC sample to examine a causal association from psychotic experiences to cognitive function.

## Results

### Psychotic experiences

At age 18, PLIKSi data were available from 4,718 participants, of whom 374 participants (8%) reported PLEs within the last six months (n=176 suspected, n=198 definite). At age 24, PLIKSi data were available from 3,889 participants, of whom 409 participants (11%) reported PLEs within the last year (n=153 suspected, n=256 definite). For the 2,822 participants with PLIKSi data available across both measurement occasions, PLEs remitted between age 18 and 24 for 130 participants, arose at age 24 where not present at age 18 for 174 participants, and persisted across both measurement occasions for 85 participants. Anxiety and depressive disorders were common in individuals who experience PLEs. Demographic and comorbidity data are provided in Supplementary Table S1. An overview of cognitive data across groups is provided in Supplementary Figure S2.

### Observational analyses

For studies 1 and 2, we primarily report the findings from the multiply imputed datasets, due to the greater potential for biased estimates in complete case analyses^28^. Complete case models for both studies are reported in the Supplementary Material.

#### Study 1

There was strong evidence for a cross-sectional association between PLEs and poorer working memory at age 24, and this was robust to all levels of adjustment (Table 1, see Table S2 for complete case analyses). There was also evidence for an association between PLEs and poorer response inhibition (Table 1). Evidence for this association was weaker in the complete case analysis; however, effect estimates were consistent (i.e., in the same direction) across all models. We did not find clear evidence to suggest a cross-sectional association between emotion recognition performance and PLEs in any analyses (Tables 1 and 2, Supplementary Tables S2 and S3).

**Table 1.**
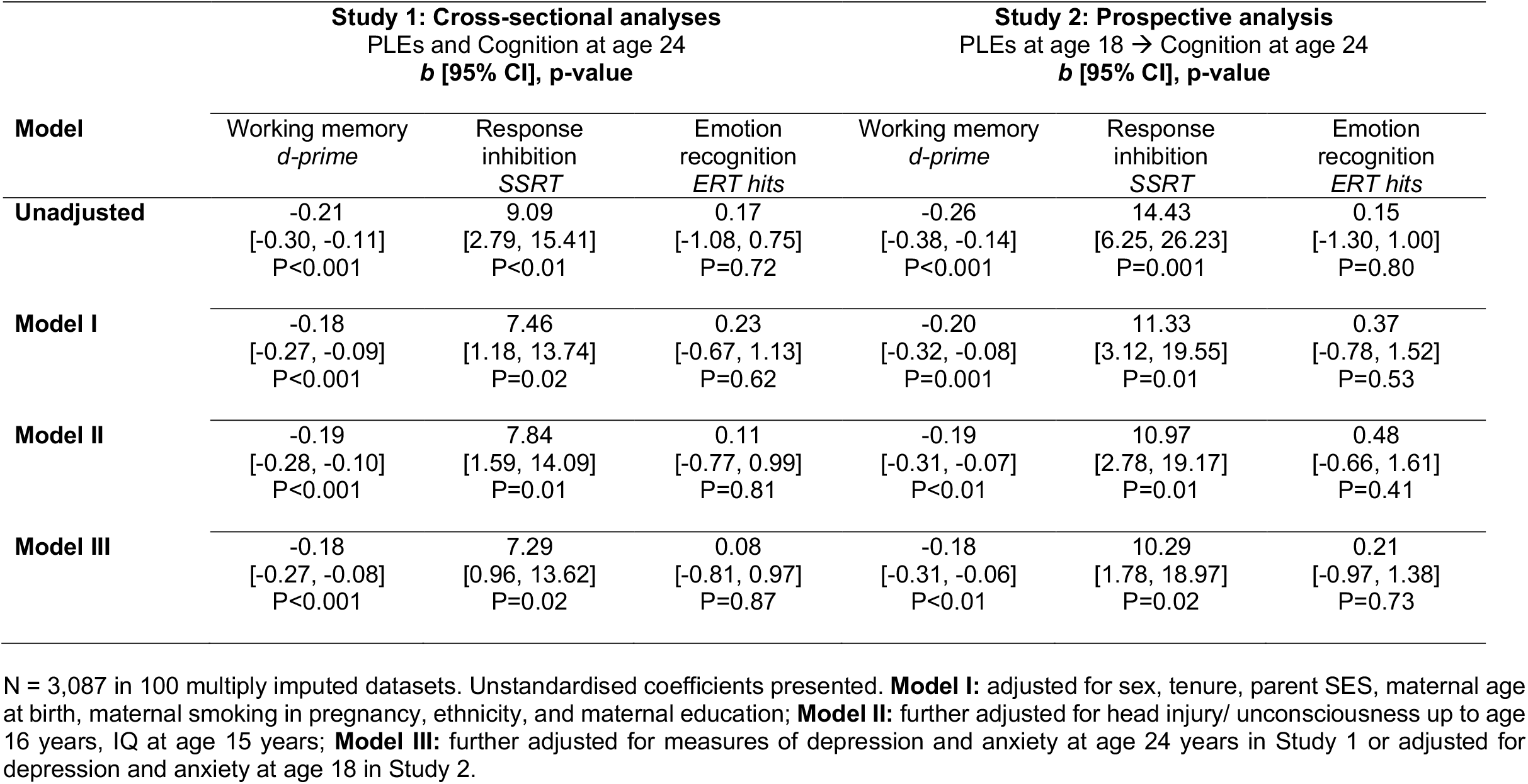
Associations between psychosis-like symptoms and cognitive functioning: cross-sectional and prospective analyses.

|  | **Study 1: Cross-sectional analyses**<br>PLEs and Cognition at age 24<br>*b* [95% CI], *p*-value |  |  | **Study 2: Prospective analysis**<br>PLEs at age 18 → Cognition at age 24<br>*b* [95% CI], *p*-value |  |  |
| --- | --- | --- | --- | --- | --- | --- |
|  | Working memory<br>*d-prime* | Response<br>inhibition<br>*SSRT* | Emotion<br>recognition<br>*ERT hits* | Working memory<br>*d-prime* | Response<br>inhibition<br>*SSRT* | Emotion<br>recognition<br>*ERT hits* |
| **Unadjusted** | -0.21<br>[-0.30, -0.11]<br>P<0.001 | 9.09<br>[2.79, 15.41]<br>P<0.01 | 0.17<br>[-1.08, 0.75]<br>P=0.72 | -0.26<br>[-0.38, -0.14]<br>P<0.001 | 14.43<br>[6.25, 26.23]<br>P=0.001 | 0.15<br>[-1.30, 1.00]<br>P=0.80 |
| **Model I** | -0.18<br>[-0.27, -0.09]<br>P<0.001 | 7.46<br>[1.18, 13.74]<br>P=0.02 | 0.23<br>[-0.67, 1.13]<br>P=0.62 | -0.20<br>[-0.32, -0.08]<br>P=0.001 | 11.33<br>[3.12, 19.55]<br>P=0.01 | 0.37<br>[-0.78, 1.52]<br>P=0.53 |
| **Model II** | -0.19<br>[-0.28, -0.10]<br>P<0.001 | 7.84<br>[1.59, 14.09]<br>P=0.01 | 0.11<br>[-0.77, 0.99]<br>P=0.81 | -0.19<br>[-0.31, -0.07]<br>P<0.01 | 10.97<br>[2.78, 19.17]<br>P=0.01 | 0.48<br>[-0.66, 1.61]<br>P=0.41 |
| **Model III** | -0.18<br>[-0.27, -0.08]<br>P<0.001 | 7.29<br>[0.96, 13.62]<br>P=0.02 | 0.08<br>[-0.81, 0.97]<br>P=0.87 | -0.18<br>[-0.31, -0.06]<br>P<0.01 | 10.29<br>[1.78, 18.97]<br>P=0.02 | 0.21<br>[-0.97, 1.38]<br>P=0.73 |

N = 3,087 in 100 multiply imputed datasets. Unstandardised coefficients presented. **Model I:** adjusted for sex, tenure, parent SES, maternal age at birth, maternal smoking in pregnancy, ethnicity, and maternal education; **Model II:** further adjusted for head injury/ unconsciousness up to age 16 years, IQ at age 15 years; **Model III:** further adjusted for measures of depression and anxiety at age 24 years in Study 1 or adjusted for depression and anxiety at age 18 in Study 2.

**Table 2.** Associations between psychosis-like symptoms and emotion-specific hit rate on the Emotion Recognition Task: cross-sectional and prospective analyses.

|  |  | Emotion Specific Hit Rate (*b* [95% CI]) |  |  |  |  |  | Wald test |
| --- | --- | --- | --- | --- | --- | --- | --- | --- |
|  |  | Anger | Disgust | Fear | Happy | Sad | Surprise | Statistic,<br>p-value |
| Study 1:<br>Cross-sectional | Unadjusted | 0.02<br>[-0.27, 0.32] | 0.07<br>[-0.20, 0.34] | -0.01<br>[-0.41, 0.40] | -0.15<br>[-0.40, 0.11] | -0.05<br>[-0.30, 0.21] | -0.06<br>[-0.23, 0.12] | 0.43,<br>P=0.86 |
|  | Model I | 0.11<br>[-0.18, 0.40] | 0.14<br>[-0.13, 0.41] | 0.12<br>[-0.29, 0.52] | -0.15<br>[-0.40, 0.11] | 0.03<br>[-0.22, 0.28] | -0.01<br>[-0.20, 0.17] | 0.53,<br>P=0.79 |
|  | Model II | 0.08<br>[-0.21, 0.36] | 0.12<br>[-0.15, 0.39] | 0.08<br>[-0.32, 0.48] | -0.15<br>[-0.40, 0.11] | -0.00<br>[-0.25, 0.25] | -0.02<br>[-0.20, 0.16] | 0.46,<br>P=0.84 |
|  | Model III | 0.08<br>[-0.21, 0.37] | 0.10<br>[-0.17, 0.37] | 0.05<br>[-0.36, 0.45] | -0.09<br>[-0.34, 0.17] | -0.06<br>[-0.31, 0.19] | -0.00<br>[-0.18, 0.19] | 0.32,<br>P=0.93 |
| Study 2: Prospective<br>PLEs → Cognition | Unadjusted | 0.19<br>[-0.18, 0.55] | 0.25<br>[-0.08, 0.58] | -0.18<br>[-0.69, 0.32] | -0.06<br>[-0.38, 0.27] | -0.19<br>[-0.50, 0.14] | -0.17<br>[-0.40, 0.07] | 1.58,<br>P=0.15 |
|  | Model I | 0.30<br>[-0.07, 0.67] | 0.33<br>[-0.01, 0.66] | 0.05<br>[-0.46, 0.55] | -0.13<br>[-0.45, 0.20] | -0.06<br>[-0.39, 0.26] | -0.13<br>[-0.36, 0.11] | 1.62,<br>P=0.14 |
|  | Model II | 0.34<br>[-0.03, 0.70] | 0.35<br>[0.01, 0.68] | 0.09<br>[-0.43, 0.59] | -0.12<br>[-0.45, 0.20] | -0.04<br>[-0.36, 0.28] | -0.12<br>[-0.35, 0.11] | 1.69,<br>P=0.12 |
|  | Model III | 0.32<br>[-0.06, 0.70] | 0.24<br>[0.11, 0.59] | 0.07<br>[-0.57, 0.46] | -0.07<br>[-0.41, 0.27] | -0.07<br>[-0.40, 0.27] | -0.15<br>[-0.39, 0.10] | 1.30,<br>P=0.25 |

N = 3,087 in 100 multiply imputed datasets. Unstandardised coefficients presented. **Model I:** adjusted for sex, tenure, parent SES, maternal age at birth, maternal smoking in pregnancy, ethnicity, and maternal education; **Model II:** further adjusted for head injury/ unconsciousness up to age 16 years, IQ at age 15 years; **Model III:** further adjusted for measures of depression and anxiety at age 24 years in Study 1 or adjusted for depression and anxiety at age 18 in Study 2.

#### Study 2

Our results indicated that participants reporting PLEs at age 18 experience poorer working memory at age 24 (Table 1), with consistent results in the complete case analysis (Supplementary Table S2). Examining the prospective association in the reverse direction, from working memory at age 18 to PLEs at 24, we found no evidence for an association across all levels of adjustments (ORs 0.93 to 0.99; Supplementary Table S4).

There was evidence for a prospective association between PLEs at age 18 and poorer response inhibition at age 24, robust to all levels of adjustment (Table 1). Slightly weaker evidence for this association was also found in the complete case analysis (Supplementary Table S2). We found no clear evidence to suggest a prospective association between global or specific emotion recognition and PLEs (Tables 1 and 2), with comparable results in complete cases (Supplementary Tables S2 and S3).

### Mendelian randomization analyses

#### Study 3

Results from two-sample MR analyses were ambiguous (Table 3), most likely because they were underpowered. We found a consistent (negative) direction of effect estimates across all four methods for emotion recognition, but the confidence intervals for these estimates were wide. We found a mixed direction of effect estimates across MR methods for working memory and response inhibition, with similar imprecision.

**Table 3.**
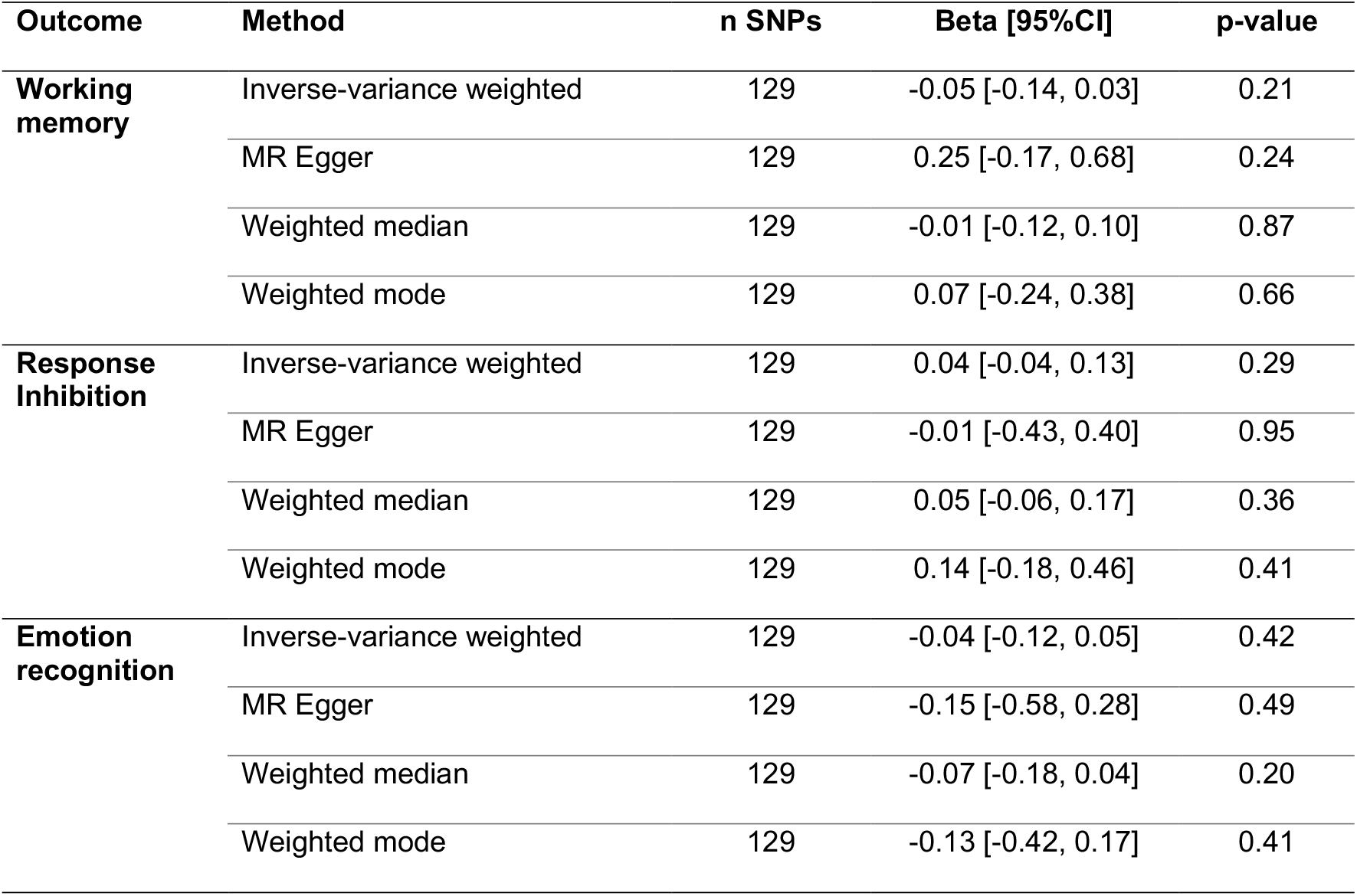
Two sample Mendelian randomisation analysis of the effect of genetic liability to schizophrenia on cognitive function.

For one-sample MR analyses, polygenic risk scores for schizophrenia did not predict PLEs within our sample (Estimate= 23.22, 95% CI −56.13 to 102.98, p=.567). We therefore did not progress these analyses beyond the validation stage.

## Discussion

We examined cross-sectional and prospective relationships between experiences of psychosis and specific cognitive domains (working memory, response inhibition and emotion recognition) in a population sample. We found evidence of both cross-sectional and prospective associations between PLEs and poorer cognitive test performance, in particular in the domains of working memory and response inhibition, indicating that PLEs at age 18 predict poorer later cognitive function in these domains over a period of 6 years. However, MR analyses did not provide evidence to allow us to conclude that these associations reflect causal pathways.

### Comparison with previous studies

Our findings extend previous research showing cross-sectional associations between PLEs and impairments in specific domains of cognitive function, including working memory^10^. By using a longitudinal approach, we were able to evaluate associations over a key developmental stage of cognitive maturation^13^, and vulnerability for development of PLEs^2^ and psychotic disorders^14^.

Previous research has examined longitudinal associations between general cognitive function, as measured by IQ, and PLEs early in childhood and adolescence, showing that lower IQ is associated with a higher rate of later PLEs^12^. Here we find rather that PLEs predict poorer performance in specific domains of cognitive function when assessed later in life. This may indicate bi-directional causal pathways of PLEs with cognitive function.

The longitudinal nature of ALSPAC data allows us to adjust for key confounders, including socioeconomic and gestational factors, head injury and earlier global cognitive function. PLEs are also associated with affective and anxiety disorders^1^, which themselves are linked to cognitive impairment^37,38^. Therefore, adjusting for depression and anxiety is important to identify cognitive impairments specifically associated with PLEs, rather than simply those linked to co-occurring mental health problems. Results robust to these adjustments improves confidence that the associations we report are not the result of concurrent or historical confounders.

MR analyses did not provide evidence to allow us to conclude that the associations we observed reflect causal pathways. Previous research has shown negative genetic associations between general cognition and schizophrenia using a polygenic risk score (PRS) approach^39^, and that high IQ attenuates likelihood of schizophrenia in research using an MR approach^40^. Conducting MR analyses to identify causal pathways from specific cognitive domains, such as response inhibition and working memory, may identify targets for intervention. Unfortunately, our MR analyses were most likely underpowered, since GWAS of individual cognitive domains did not yield any genome-wide significant results. Moreover, we found that the PRS for schizophrenia did not predict PLEs at age 24, limiting our ability to complete a one-sample MR analysis. This result is in keeping with prior research in ALSPAC showing that genetic liability for schizophrenia is associated primarily with negative symptoms rather than with PLEs^41,42^. Therefore, MR analyses of causal pathways between specific cognitive functions and PLEs remain an important area for future research, should cognitive GWAS of suitable sample sizes become available.

### Limitations

There are a number of limitations to this study. First, it is possible that the causal pathway could be in the other direction to that tested by our MR analysis; that is, that liability for cognitive impairment may increase the risk of PLEs. Due to lack of robust instruments for our measures of cognition, MR was only completed with schizophrenia as exposure to cognition as outcome. Second, ALSPAC suffers from attrition, with lower participation amongst socially disadvantaged and lesser educated participants^43^. Polygenic risk scores for schizophrenia are associated with drop-out, which can lead to underestimation of risk of schizophrenia and related psychiatric and behavioural phenotypes^44^. However, potential bias arising from missing data was dealt with using multiple imputation, using a large amount of additional information to make the assumption of missing at random as plausible as possible. Findings from complete case and multiple imputed models were comparable for each of the three outcomes. Third, we could not complete bidirectional prospective analyses for all cognitive measures, since equivalent data for response inhibition and emotion recognition were not available at age 18. Furthermore, the sample at age 24 has not yet passed through the entire risk period for PLEs^2,14^, indicating that further longitudinal research, with repetition of cognitive and PLE assessments is required to shed light on the developmental trajectories of cognition and PLEs and their association in later adulthood.

### Implications and conclusions

Young people who experience PLEs are likely to also experience other mental health and functional impairments. Here we show that these impairments in young adults with PLEs extend to concurrent and future poorer function in the cognitive domains of working memory and response inhibition.

## Data Availability

Any researcher can apply to use ALSPAC data, including the variables under investigation in this study. Access information is provided here: http://www.bristol.ac.uk/alspac/researchers/access/

## Acknowledgements

We are extremely grateful to all the families who took part in this study, the midwives for their help in recruiting them, and the whole ALSPAC team, which includes interviewers, computer and laboratory technicians, clerical workers, research scientists, volunteers, managers, receptionists and nurses.

