## Supplementary Material for "Psychosis-like experiences and cognition in young adults: an observational and Mendelian randomisation study"

**Supplementary Figure S1.** A timeline of data used in analyses.

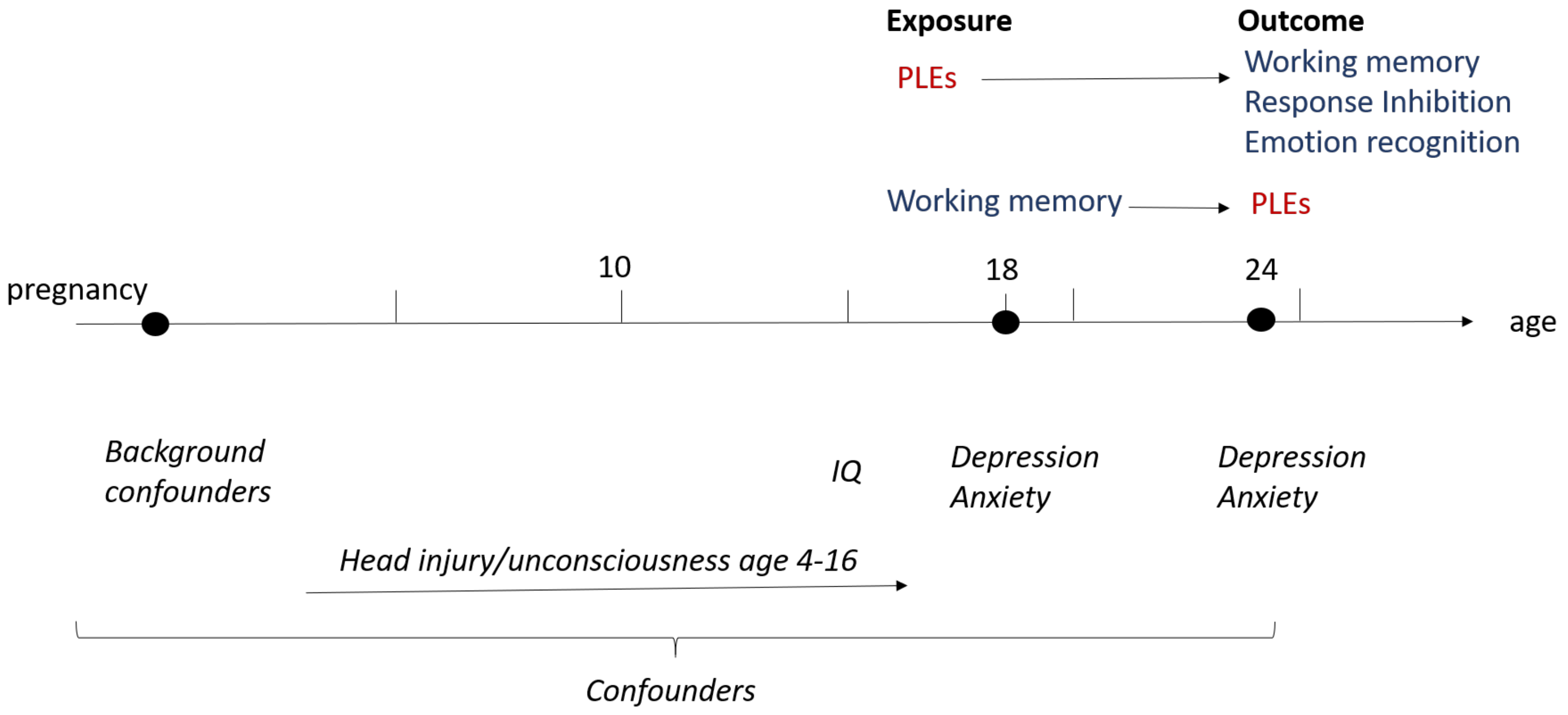


Confounders controlled for in analyses are italicized. PLEs: Psychosis-like experiences measured with the PLIKSi, IQ: Intelligence Quotient measured using the Wechsler Abbreviated Scales of Intelligence.

**Supplementary Figure S2.** Boxplots of cross-sectional association of PLEs at age 24 and cognitive function at 24 (left column) and prospective association of PLEs at age 18 and cognitive function at 24 (right column).

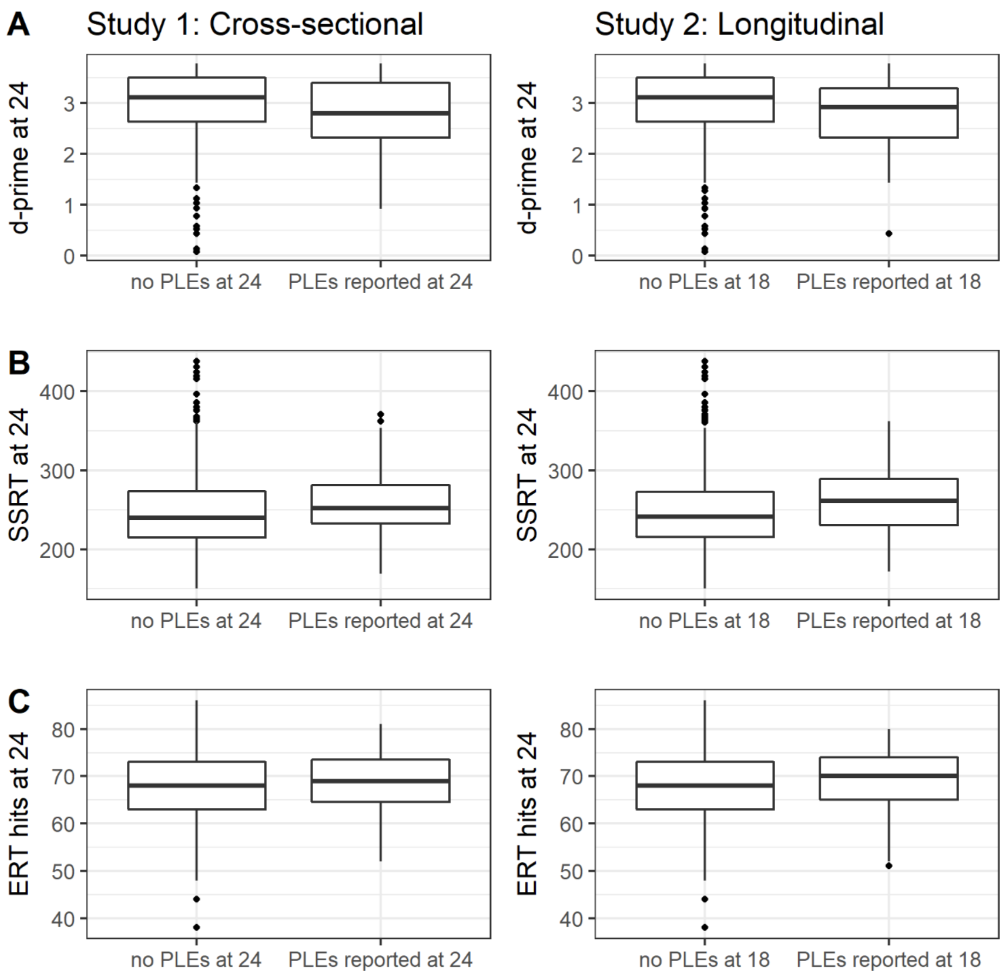


A: working memory, B: response Inhibition, C: facial emotion recognition.

**Supplementary Table S1.** Demographic and comorbidity information for study sample.

| Age | PLEs reported | N | N Male (%) | N Depressed (%) | N Anxiety disorder (%) |
|---|---|---|---|---|---|
| 18 | Suspected/Definite | 374 | 134 (36%) | 78 (21%) | 97 (26%) |
| | None | 4,343 | 1,917 (44%) | 257 (6%) | 291 (7%) |
| 24 | Suspected/Definite | 409 | 159 (39%) | 94 (23%) | 113 (28%) |
| | None | 3,480 | 1,300 (37%) | 310 (9%) | 396 (11%) |

PLEs: Psychosis-like experiences.

**Supplementary Table S2.** Associations between psychosis-like symptoms and cognitive functioning: cross-sectional and prospective analyses.

| | **Study 1: Cross-sectional analyses (n=2,134)** PLEs and Cognition at age 24 ***b* [95% CI], P-value** | | | **Study 2: Prospective analysis (n=1,788)** PLEs at age 18 → Cognition at age 24 ***b* [95% CI], P-value** | | |
|---|---|---|---|---|---|---|
| **Model** | Working memory *d-prime* | Response inhibition *SSRT* | Emotion Recognition *ERT hits* | Working memory *d-prime* | Response inhibition *SSRT* | Emotion Recognition *ERT hits* |
| **Unadjusted** | -0.23 [-0.34, -0.11] P<0.001 | 6.47 [-1.15, 14.10] P=0.10 | 0.09 [-1.03, 1.21] P=0.88 | -0.20 [-0.35, -0.06] P<0.01 | 10.89 [1.42, 20.36] P=0.02 | 0.61 [-0.80, 2.01] P=0.40 |
| **Model I** | -0.21 [-0.33, -0.10] P<0.001 | 5.61 [-2.00, 13.23] P=0.15 | 0.41 [-0.70, 1.51] P=0.47 | -0.17 [-0.32, -0.03] P=0.02 | 9.66 [0.14, 19.18] P=0.05 | 0.83 [-0.57, 2.23] P=0.24 |
| **Model II** | -0.22 [-0.34, -0.11] P<0.001 | 6.05 [-1.52, 13.63] P=0.12 | 0.25 [-0.83, 1.33] P=0.65 | -0.17 [-0.31, -0.03] P=0.02 | 9.47 [-0.01, 18.96] P=0.05 | 0.90 [-0.45, 2.25] P=0.19 |
| **Model III** | -0.22 [-0.34, -0.11] P<0.001 | 5.53 [-2.12, 13.19] P=0.16 | 0.23 [-0.86, 1.32] P=0.68 | -0.16 [-0.30, -0.02] P=0.03 | 8.75 [-0.97, 18.47] P=0.08 | 0.62 [-0.77, 2.00] P=0.38 |

Note. Unstandardised coefficients presented. **Model I:** adjusted for sex, tenure, parent SES, maternal age at birth, maternal smoking in pregnancy, ethnicity, and maternal education; **Model II:** further adjusted for head injury/ unconsciousness up to age 16 years, IQ at age 15 years; **Model III:** further adjusted for measures of depression and anxiety at age 24 years in Study 1, or adjusted for depression and anxiety at age 18 in Study 2.

**Supplementary Table S3.** Associations between psychosis-like symptoms and emotion-specific hit rate on the Emotion Recognition Task: cross-sectional and prospective analyses.

| | | **Emotion Recognition** *Emotion Specific Hit rate* ***b*** **[95% CI]** | | | | | | **Wald test Statistic, P-value** |
|---|---|---|---|---|---|---|---|---|
| **Models** | | **Anger** | **Disgust** | **Fear** | **Happy** | **Sad** | **Surprise** | |
| **Study 1: Cross-sectional (n=2,300)** | **Unadjusted** | 0.17 [-0.18, 0.52] | 0.18 [-0.15, 0.51] | 0.05 [-0.43, 0.54] | -0.11 [-0.42, 0.20] | -0.09 [-0.39, 0.22] | -0.05 [-0.27, 0.17] | 0.65, P=0.69 |
| | **Model I** | 0.25 [-0.11, 0.60] | 0.24 [-0.09, 0.56] | 0.17 [-0.31, 0.65] | -0.11 [-0.42, 0.20] | -0.03 [-0.33, 0.28] | -0.01 [-0.23, 0.21] | 0.82, P=0.55 |
| | **Model II** | 0.21 [-0.14, 0.55] | 0.21 [-0.11, 0.54] | 0.12 [-0.36, 0.59] | -0.12 [-0.42, 0.19] | -0.06 [-0.36, 0.24] | -0.02 [-0.24, 0.20] | 0.72, P=0.63 |
| | **Model III** | 0.21 [-0.13, 0.55] | 0.20 [-0.13, 0.52] | 0.09 [-0.39, 0.56] | -0.05 [-0.36, 0.26] | -0.11 [-0.41, 0.19] | -0.01 [-0.23, 0.21] | 0.67, P=0.67 |
| **Study 2: Prospective (n=1,918)** | **Unadjusted** | 0.17 [-0.27, 0.61] | 0.35 [-0.06, 0.76] | 0.14 [-0.45, 0.74] | -0.06 [-0.43, 0.32] | -0.14 [-0.51, 0.23] | -0.06 [-0.32, 0.21] | 0.88, P=0.51 |
| | **Model I** | 0.25 [-0.19, 0.69] | 0.37 [-0.05, 0.78] | 0.24 [-0.35, 0.84] | -0.13 [-0.50, 0.25] | -0.08 [-0.45, 0.29] | -0.04 [-0.31, 0.23] | 0.99, P=0.43 |
| | **Model II** | 0.27 [-0.16, 0.70] | 0.38 [-0.03, 0.78] | 0.27 [-0.32, 0.86] | -0.13 [-0.50, 0.25] | -0.06 [-0.43, 0.30] | -0.04 [-0.31, 0.23] | 1.06, P=0.39 |
| | **Model III** | 0.23 [-0.21, 0.66] | 0.28 [-0.14, 0.69] | 0.10 [-0.50, 0.70] | -0.08 [-0.47, 0.31] | -0.07 [-0.44, 0.31] | -0.04 [-0.31, 0.23] | 0.59, P=0.74 |

Note. Complete cases analysis. Unstandardised coefficients presented. **Model I:** adjusted for sex, tenure, parent SES, maternal age at birth, maternal smoking in pregnancy, ethnicity, and maternal education; **Model II:** further adjusted for head injury/ unconsciousness up to age 16 years, IQ at age 15 years; **Model III:** further adjusted for measures of depression and anxiety at 24 years of age in Study 1, or adjusted for depression and anxiety at age 18 for Study 2.

**Supplementary Table S4.** Associations between working memory at age 18 years and psychotic-like symptoms at age 24 years (unstandardized coefficients).

| | **Complete case (n=2,134)** Working memory at age 18 → PLEs age 24 | | **Multiple imputation (n=3,087)** Working memory at age 18 → PLEs age 24 | |
|---|---|---|---|---|
| | **OR [95% CI]** | **P-value** | **OR [95% CI]** | **P-value** |
| **Unadjusted** | 0.97 [0.81, 1.16] | 0.74 | 0.93 [0.81, 1.06] | 0.27 |
| **Model I** | 1.00 [0.83, 1.20] | 0.97 | 0.97 [0.85, 1.12] | 0.71 |
| **Model II** | 1.01 [0.83, 1.21] | 0.99 | 0.97 [0.84, 1.12] | 0.66 |
| **Model III** | 1.00 [0.82, 1.23] | 0.96 | 0.99 [0.85, 1.15] | 0.91 |

Note. **Model I:** adjusted for sex, tenure, parent SES, maternal age at birth, maternal smoking in pregnancy, ethnicity, and maternal education; **Model II:** further adjusted for head injury/ unconsciousness up to age 16 years, IQ at age 15 years; **Model III:** further adjusted for measures of depression and anxiety at age 18.
